# A longitudinal comparison of spike and nucleocapsid SARS-CoV-2 antibody responses in a tertiary hospital’s laboratory workers with validation of DBS specimen analysis

**DOI:** 10.1101/2020.10.29.20219931

**Authors:** I Murrell, D Forde, L Tyson, L Chichester, A Garratt, O Vineall, N Palmer, R Jones, C Moore

**Author notes:** Co-first authorship (I Murrell, D Forde).

## Abstract

There is a requirement for easily accessible, high throughput serological testing as part of the SARS-CoV-2 pandemic response. Whilst of limited diagnostic use in an acute individual setting, its use on a population level is key to informing a coherent public health response. As experience of commercial assays increases, so too does knowledge of their precision and limitations. Here we present our experience of these systems thus far. We perform a spot sero-prevalence study amongst staff in a tertiary hospital’s clinical microbiology laboratory, before undertaking validation of DBS serological testing as an alternate specimen for analysis. Finally, we characterise the spike and nucleocapsid antibody response over 160 days post a positive PCR test in nine non-hospitalised staff members.

Amongst a cohort of 195 staff, 17 tested positive for SARS-CoV-2 antibodies (8.7%). Self-reporting of SARS-CoV2 infection (P=<0.0001) and testing of a household contact (P = 0.027) were significant variables amongst the positive and negative sub-groups. Testing of 28 matched serum and DBS samples demonstrated 96% accuracy between the sample types. A differential rate of decline of SARS-CoV-2 antibodies against nucleocapsid or spike protein was observed. At 4 months post a positive PCR test 7/9 (78%) individuals had detectable antibodies against spike protein, but only 2/9 (22%) had detectable antibodies against nucleocapsid protein. This study reveals a broad agreement amongst commercial platforms tested and suggests the use of DBS as an alternate specimen option to enable widespread population testing for SARS-CoV-2 antibodies. These results suggest potential limitations of these platforms in estimating historical infection. By setting this temporal point of reference for this cohort of non-patient facing laboratory staff, future exposure risks and mitigation strategies can be evaluated more fully.

## 1 Introduction

Severe Acute Respiratory Syndrome Coronavirus 2 (SARS-CoV-2) is the 7^th^ coronavirus to infect humans, following the relatively recent discoveries of SARS-CoV-1, MERS-CoV, hCoV-NL63 and hCoV-HKU1(Drosten et al., 2003; Ksiazek et al., 2003; Van Der Hoek et al., 2004; Woo et al., 2005; Zaki et al., 2012). First identified as the causative agent of a cluster of pneumonia cases in late December 2019 in Hubei province, China, SARS-CoV-2 rapidly spread globally and was declared a pandemic on the 11^th^ March 2020. As of 25th October 2020 there have been a documented 42,512,186 cases and 1,147,301 deaths reported globally to WHO (WHO, 2020).

The detection of viral genome using molecular techniques remains the gold standard for the laboratory diagnosis of patients presenting with acute SARS-CoV-2 infection. However, a successful response to the pandemic also requires the complementary use of serological testing to accurately identify the proportion of the population exposed and infected by SARS-CoV-2 with and without discernible clinical symptoms. The data generated from such sero-prevalence studies will inform future public health strategies by enabling the analysis of exposure risk, disease severity and by determining the size of the remaining susceptible population (Infantino et al., 2020; Theel et al., 2020; Weitz et al., 2020).

Commercial serological assays have been quickly developed with the capabilities to detect antibodies to either the spike protein and/or the nucleocapsid protein. However, validation data and nuances regarding the kinetics, duration and potential neutralisation effects of the detected antibody are only beginning to emerge (Favresse et al., 2020; Kohmer et al., 2020; Krüttgen et al., 2020; Ma et al., 2009; Perreault et al., 2020; Tang et al., 2020; Wang et al., 2020; Yongchen et al., 2020). Most serological assays are only validated for the use of serum or plasma that limits their use for high throughput mass population testing, due in part to the logistical constraints of collecting venous blood samples. To facilitate mass population screening alternative sample types such as dry blood spots (DBS) are attractive, as the use of this specimen type has clear benefits logistically and in terms of economic allocation of precious healthcare resources.

## 2 Objectives

- To validate four commercially available assays for the detection of antibody to SARS-CoV-2 using a single panel made up of serum samples collected prior to December 2019, and a panel of serum samples from laboratory confirmed COVID19 cases.
- To perform a point sero-prevalence study in the clinical microbiology laboratory of Public Health Wales to determine the past infection rate of a largely non-patient facing staff group and to determine if a perceived history of past infection with SARS-CoV-2 was confirmed by serology.
- To validate the use of DBS samples for SARS-CoV-2 serology using a commercial platform.
- To characterise the longitudinal antibody response against the nucleocapsid and spike proteins in a non-hospitalised cohort.

## 3 Materials and Methods

### 3.1 Composition of serum panel for assay validation

To test the assay specificity, 60 stored serum samples collected prior to December 2019 were selected from the antenatal/blood born virus screening bio-bank. In addition, 10 additional samples were included from patients who tested positive to one of the seasonal coronaviruses (NL63 n=3, HKU1 n=2, OC43 n=4, 229E n=1) and negative for SARS-CoV-2 by RT-PCR. To determine sensitivity, serum from 41 laboratory-confirmed, hospitalised COVID19 cases were collected. All samples were collected at a minimum of 14 days after the initial positive molecular result.

This panel was tested across four enzyme immunoassay (EIA) platforms following the manufacturers’ instructions. Three were performed on random access high-throughput platforms; the Anti-SARS-CoV-2 total antibody assay (Ortho Clinical Diagnostics) and the Anti-SARS-CoV-2 IgG assay (Ortho Clinical Diagnostics) which both target the spike protein were performed on the Vitros 3600 and the Abbott SARS-CoV-2 IgG assay (Abbott) which targets the nucleocapsid protein which was performed on the Abbott Architect. The fourth assay; was a plate based, semi-quantitative SARS-CoV-2 IgG ELISA by Euroimmun targeting the spike protein and was performed using the automated DS2 platform (Dynex).

### 3.2 Participants in the point sero-prevalence study

All staff from administrative, scientific, non-patient facing medical staff and support divisions in the Microbiology laboratory of Public Health Wales, Cardiff were invited to participate by internal email. A questionnaire was completed upon enrolment and phlebotomy undertaken across a five-day period commencing 8^th^ June 2020. All samples were tested using the Euroimmun assay. The local ethics board was consulted prior to sampling.

### 3.3 Dried blood spot testing

Participants from the sero-prevalence study who returned a positive antibody result were invited to submit a DBS sample, this was collected within seven days of the initial serum collection. A matched number of negative controls from the same cohort were also obtained. DBS samples were collected by a single healthcare provider using standard collection procedure (Edelbroek et al., 2009). Briefly a fingerprick sample of blood was collected using lancet (Unistik 3, Owen Mumford) and three blood spots targets completed on a cellulose based card (Whatman 903, Cytiva). These were allowed to dry at room temperature before being moved for refrigerated storage. The sera underwent repeat manual analysis in tandem with their corresponding DBS samples for comparison. Briefly, two DBS spots were submersed in 800μL of 0.05% PBS/Tween and agitated using a shaker at 700 rpm for 30mins at room temperature. Samples were refrigerated overnight. The following day the agitation was repeated, before eluates were collected to fresh storage tubes. Both serum and DBS elute were tested in triplicate using the SARS-CoV-2 IgG ELISA, (Euroimmun) assay according to manufacturer’s instructions for manual analysis. Where described, samples were diluted with sample buffer from the kit.

### 3.4 Longitudinal sero-study

Participants that were identified as having a detectable SARS-CoV-2 RNA in a throat swab by RT-PCR were invited to submit monthly blood samples. These participants were all staff of Public Health Wales, Cardiff. Nine staff members were enrolled - none of whom needed hospitalisation for their initial SARS-CoV-2 infection. Samples were collected and stored over a four month period before analysis took place. The antibody response of both nucleocapsid and spike protein was evaluated by using the SARS-CoV-2 IgG assay by Abbott and the SARS-CoV-2 IgG ELISA by Euroimmun respectively.

### 3.5 Statistical analysis

Categorical variables are presented as numbers and percentages. To compare the sub-groups in the sero-study two-tailed Chi-squared and Fishers t-tests are applied as appropriate. Statistical analysis was performed using Prism V7 where a P value of <0.05 is considered significant.

## 4 Results

### 4.1 Sensitivity and specificity of four commercial SARS-CoV-2 EIA assays

The sensitivity for all four assays was greater than 85%, with each demonstrating a specificity of 100%. None of the samples from the seasonal coronavirus positive cohort showed reactivity for SARS-CoV-2 in any of the assays being validated.

Individual assay sensitivity results were; Vitros Anti-SARS-CoV-2 Total (Ortho Clinical Diagnostics) detected 39/41 of positive samples (sensitivity 95%); Vitros Anti-SARS-CoV-2 IgG (Ortho Clinical Diagnostics) detected 34/38 of positive samples (sensitivity 89%); SARS-CoV-2 IgG (Abbott) detected 37/41 of positive samples (sensitivity 90%) and the SARS-CoV-2 IgG ELISA (Euroimmun) detected 36/41 (sensitivity 88%). There was a single patient with SARS-CoV-2 detected by RT-PCR on a throat swab who had a clinically compatible presentation that was serologically negative across all assays.

### 4.2 Point sero-prevalence of SARS-CoV-2 in laboratory staff

Of the 195 individuals tested, 174 tested negative, 17 tested positive, and 4 were in the equivocal range of the assay (Table 1). Two of the four equivocal samples had repeat serology one month later and remained equivocal. This yields a point sero-positivity of 17/195 (8.7%).

**Table 1:** Baseline demographics and sub-group analysis of laboratory staff in a point SARS-CoV-2 sero-prevalence study.

|  | Total population<br>(n=195) | SARS2-CoV-2 test result |  | P value |
| --- | --- | --- | --- | --- |
|  |  | Positive<br>(n=17) | Negative<br>(n=174) |  |
| Gender – N (%) |  |  |  | .296 |
| Female | 117 (60) | 8 (47) | 109 (63) |  |
| Male | 78 (40) | 9 (53) | 65 (37) |  |
| Age – N (%) |  |  |  | .108 |
| ≤ 30y | 67 (34) | 9 (53) | 57 (33) |  |
| 31-50y | 102 (52) | 8 (47) | 91 (52) |  |
| ≥ 51y | 26 (13) | - | 26 (15) |  |
| Total No in household – N (%) |  |  |  | .141 |
| Sole occupant | 35 (18) | 2 (12) | 33 (19) |  |
| > 1 occupant | 160 (82) | 15 (88) | 141 (81) |  |
| > 2 occupants | 97 (50) | 10 (59) | 85 (49) |  |
| > 3 occupants | 62 (32) | 8 (47) | 53 (30) |  |
| > 4 occupants | 23 (12) | 6 (35) | 17 (10) |  |
| Additional keyworker in household – N (%) |  |  |  | .074 |
| No | 99 (51) | 5 (29) | 94 (54) |  |
| Yes | 96 (49) | 12 (71) | 80 (46) |  |
| Previously tested (PCR) for SARS-CoV-2 – N (%) |  |  |  | - |
| No | 145 (74) | 7 (41) | 136 (78) |  |
| Yes – Positive | 9 (5) | 8 (47) | - |  |
| Yes – Negative | 41 (21) | 2 (12) | 38 (22) |  |
| Another household member tested (PCR) for SARS-CoV2 – N (%) |  |  |  | .027 |
| No | 165 (85) | 11 (65) | 151 (87) |  |
| Yes – Positive | 9 (5) | 5 (29) | 4 (2) |  |
| Yes – Negative | 21 (10) | 1 (6) | 19 (11) |  |
| Self-reported SARS-CoV-2 infection – N (%) |  |  |  | .0001 |
| No | 138 (71) | 3 (18) | 132 (76) |  |
| Yes | 57 (29) | 14 (82) | 42 (24) |  |
| Symptoms reported (n=57) |  |  |  |  |
| Headache | 43 (75) | 10 (59) | 31 (78) | .2 |
| Myalgia | 39 (68) | 10 (59) | 27 (68) | .557 |
| Pharyngitis | 39 (68) | 9 (53) | 28 (70) | .24 |
| Cough | 34 (60) | 8 (47) | 24 (60) | .397 |
| Fever | 25 (44) | 7 (41) | 17 (43) | .999 |
| Coryza | 22 (39) | 4 (24) | 17 (43) | .235 |
| Dyspnea | 20 (35) | 6 (35) | 13 (33) | .999 |
| Ageusia | 19 (33) | 7 (41) | 10 (25) | .342 |
| Anosmia | 18 (32) | 10 (59) | 6 (15) | .002 |
| Health condition whereby annual influenza vaccination is recommended – N (%) |  |  |  | .99 |
| No | 173 (89) | 15 (88) | 155 (89) |  |
| Yes | 22 (11) | 2 (12) | 19 (11) |  |

None of the sero-prevalence cohort had a direct patient facing role and social distancing measures were practiced within the workplace where possible. Staff who reported symptoms at any time were offered testing and isolated at home until results were available, thus reducing potential exposure in the workplace.

Only 4 of the 17 staff with positive serology participate in workplace duties that involve manipulation of diagnostic respiratory samples for SARS-CoV-2. This suggests low risk of occupational transmission under appropriate laboratory practice (CDC, n.d.; Iwen et al., 2020; WHO, n.d.).

Reassuringly, 38 of the 174 staff with negative serology had a documented negative molecular test from a throat swab collected when symptomatic.

Of the 17 staff with positive serology, 10 had a previous throat swab tested by RT-PCR for SARS-CoV-2 during a symptomatic period, of which 8 were positive. Three reported no symptoms or considered themselves not to have been infected in the past. However, after more detailed questioning post result availability, only one described a truly asymptomatic period. The remaining 2 individuals described mild coryzal symptoms that had been attributed to seasonal allergic rhinitis.

Baseline biological characteristics were similar between the positive and negative serology result groups. Two variables met statistical significance; whether a household member had met criteria to be tested for SARS-CoV-2 (Fishers test, P = 0.027) and self-reporting of likelihood of past SARS-CoV-2 (Fishers test, P = <.0001). There was a trend towards a higher percentage of household co-habitation with a keyworker in those who tested positive compared with those who tested negative (71% vs 46%). Amongst the range of symptoms reported by both groups, only anosmia reached significance (Fishers test, P = 0.003).

### 4.3 Validation of DBS as an alternate specimen sample for SARS-CoV-2 serology

It was acknowledged that the SARS-CoV-2 IgG ELISA (Euroimmun) assay has not been validated for DBS specimen analysis. Interpretation of results was as per manufacturer’s instructions for serum with a ratio optical density (OD) <0.8 defined as negative, ≥0.8 to <1.1 as borderline and ≥1.1 as positive. Local laboratory recommendations for borderline results are to repeat 4-6 weeks after initial sample. 28 DBS samples were obtained within 1 week of their matched sera collection from both positive and negative sub-groups of the laboratory staff cohort (Figure 1). There was 100% concordance between sera results produced by an automated or manual technical process. A total protocol accuracy of 96% can be achieved by a two stage process whereby initially reactive neat DBS samples are further diluted to 1:3. All 6 negative samples with false reactivity on neat analysis became negative on the second stage of testing. 1 positive sample became negative on dilution. Alternatively, a single step analysis at 1:3 dilution results in 93% sensitivity and 100% specificity. As expected, sensitivity continues to be lost upon further dilution to 1:9. It is noteworthy the similarity in positive ratio results between serum and DBS specimens processed at neat, indicating comparative equivalence if used in a semi-quantitative capacity. The increase in ratio observed in the negative neat DBS likely represents non-specific binding that is blocked upon dilution of sample with buffer from the kit.

**Figure 1:**
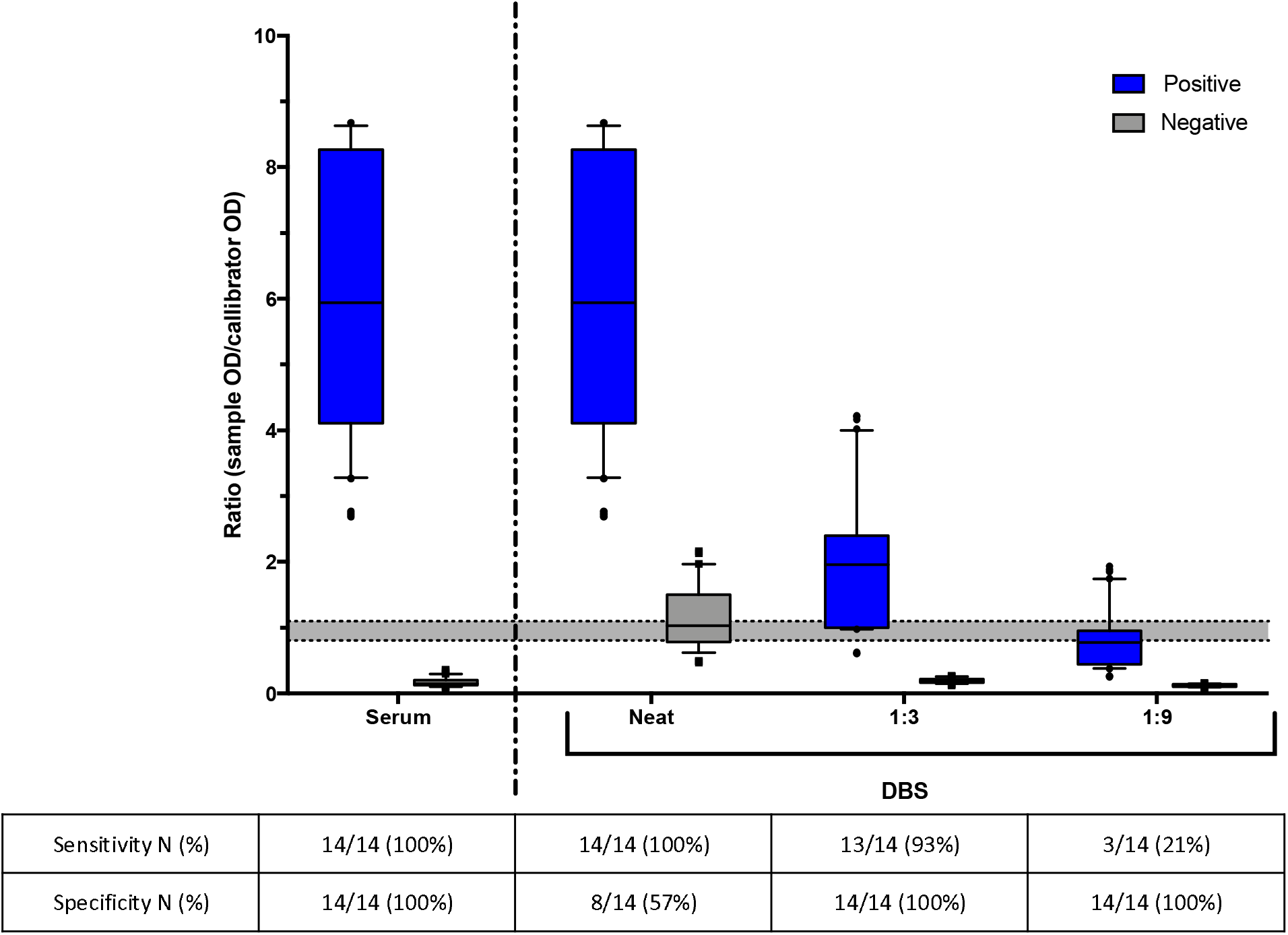
Validation of DBS specimens for SARS-CoV-2 serology. The grey shaded area represents the equivocal range of the assay, where values about this are considered positive and below are negative. Paired sera and DBS samples were collected from 28 individuals over a 7 day period. A total protocol accuracy of 96% can be achieved by a two stage process whereby initially reactive neat DBS samples are diluted to 1:3. Alternatively, a single step analysis at 1:3 dilution results in 93% sensitivity and 100% specificity. Expectedly, sensitivity decreases when further diluted to 1:9.

### 4.4 Longitudinal antibody response of SARS-CoV-2

The longitudinal antibody response to the SARS-CoV-2 spike (Figure 2A) and nucleocapsid (Figure 2B) proteins in nine non-hospitalised individuals spanning D16 to D182 post a positive PCR test is presented. The assays are semi-quantitative, where an almost unanimous decrease is observed in both antibody responses over time – one individual maintains a static spike antibody response. In the first 4 months post a positive SARS-CoV-2 PCR result on throat swab; 2/9 (22%) seroconvert towards negative spike antibody and 7/9 (78%) seroconvert towards negative nucleocapsid antibody.

**Figure 2:**
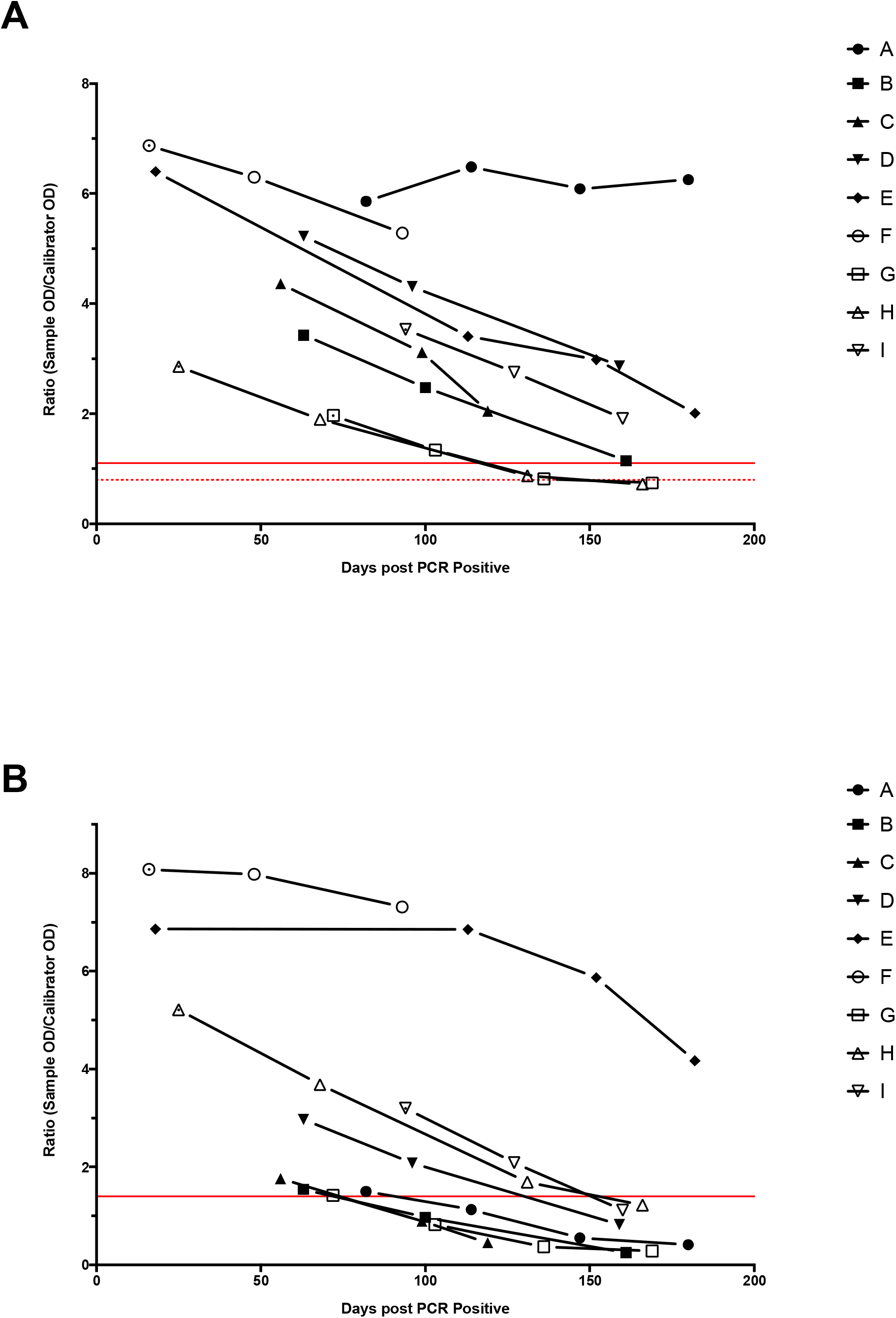
Longitudinal SARS-CoV-2 serology response in nine non-hospitalised individuals. Serum samples were collected on a monthly basis where a decrease in the positivity ratio of both assays was seen in all but one participant. Two out of nine people (22%) seroconverted their spike antibody response (A) while seven out of nine (78%) seroconverted their nucleocapsid antibody response by day 160 post a positive PCR test for SARS-CoV-2.

## 5 Discussion

There are numerous unanswered questions surrounding the use of serology for the SARS-CoV-2 pandemic (Theel et al., 2020). Accurate distinction at a population level of those that have encountered the virus from those that have not, remains the most pragmatic initial use towards an effective pandemic response. Incrementally, through populations studies this will facilitate robust answers to these initial observations; (i) that asymptomatic or milder infection results in a smaller magnitude of antibody production (Long et al., 2020; Yongchen et al., 2020), (ii) the likely duration of immunity (Deng et al., 2020; Favresse et al., 2020; Infantino et al., 2020; Perreault et al., 2020; Wang et al., 2020), (iii) that re-infection is possible (To et al., 2020; Van Elslande et al., 2020).

The validation data from our initial evaluation of four commercial assays mirrors that reported by other clinical institutions (Kohmer et al., 2020; Krüttgen et al., 2020; Tang et al., 2020). The use of a concordant sera panel facilitates robust cross-assay comparisons. All platforms tested had a sensitivity in excess of 85%, with 100% specificity. Reassuringly, there was no cross reactivity between antibodies raised to the seasonal coronaviruses in any of the assays.

Although direct human to human contact via aerosolised respiratory droplets remains the primary mode of transmission (Lai et al., 2020; Meyerowitz et al., 2020); there is growing acceptance that airborne transmission may play a minor role (Morawska and Milton, 2020; Ng et al., 2020). The SARS-CoV-2 seroprevalence rates of a populations will mirror their relative exposure risk. Therefore, patient facing healthcare workers in an aerosol generating environment will be at the highest risk of a nosocomial acquired infection - where rates of up to 12% have been observed (Lahner et al., 2020; Martin et al., 2020; Steensels et al., 2020). Clinical laboratory staff will have a comparably lower occupational exposure risk through avoidance of direct patient contact, but yet are unable to drastically modify workplace practices due to rigid working environments at a time of unprecedented workloads. Hence, they represent a valuable point of reference both on an institutional and surrounding community level. The sero-prevalence of 8.7% observed in our cohort likely reflects the presence of a lower prevalence in the surrounding community but not as high as likely to be seen in our clinical facing colleagues. Our spot sero-prevalence will also serve as a temporal benchmark to reference at a later period to estimate re-infection.

Serological DBS testing represents an invaluable tool with a proven track record in the diagnosis of HIV, Hepatitis B & C (Mohamed et al., 2013; Tuaillon et al., 2010; Uttayamakul et al., 2005). There are clear advantages in terms of sample collection/transport and economics compared to traditional phlebotomy. At a time when mass population based serological testing is a priority it represents a useful tool due to the potential for self-sampling and postal home testing kits (Parker and Cubitt, 1999; Thevis et al., 2020). Whilst not validated for DBS specimens the SARS-CoV-2 IgG ELISA by Euroimmun assay performed well with an overall test accuracy of 96% compared to sera. It is noteworthy to highlight the short period between sera and DBS sample collection and testing which in our validation was under one week. This excludes any potential bias of waning antibody with subsequent inaccurate performance data. This is reflected in the similarity in magnitude of assay signal result between the different specimens. Although performed manually, the authors see no barrier to optimise this process for automation. DBS sampling readily represents an efficient mass screening tool with options to be used as a screen to identify a reactive sub-population for verification by phlebotomy or on its own to rapidly estimate sero-prevalence rates. Both accurate and accessible SARS-CoV-2 serological diagnostics represent a crucial asset in the public health toolkit needed to curb this pandemic.

Presently, there is a gap in our knowledge about the potential differences between differing antibody targets (nucleocapsid or spike protein) regarding their sensitivity, kinetics and neutralising capabilities (Atyeo et al., 2020; Iwasaki and Yang, 2020). There is some suggestion that the spike antibody response may correlate better with neutralising antibody (Du et al., 2007; Walls et al., 2020). This is fortunate given our observation of an almost unanimous decrease in both spike and nucleocapsid antibodies over the relatively short period of four months. In fact the majority of our participants 7/9 (78%). seroconvert towards negativity in their nucleocapsid antibody response. This is particularly notable when undertaking retrospective infection exposure analysis. These first generation assays have reached the commercial forum quickly and undoubtedly can be improved upon, but they remain a valuable tool especially in light of their high specificity.

A shield immunity model (Weitz et al., 2020) coupled with a vaccine is the most likely route towards ending this pandemic. Mass serological testing could be facilitated by DBS sampling but we must be mindful of their limitations due to antibody kinetics (Thevis et al., 2020).

## 6 Conclusions

- A point seroprevalence study of laboratory staff showed a SARS-CoV-2 seropositivity rate of 8.7%.
- DBS testing a valid alternative sample type for serological analysis.
- The SARS-CoV-2 serological response wanes over time, particularly the nucleocapsid response.

## Data Availability

All data is available upon request.

## Acknowledgements

The authors wish to thank Mark Saunders, Rachel Hunt and Nigel Roberts for their assistance locating clinical samples. Thank you to the laboratory staff of Public Health Wales Microbiology for their inspirational commitment in this pandemic.

## Declarations of interes

none.

## Notes

### Competing Interest Statement

The authors have declared no competing interest.

### Funding Statement

No funding was received.

### Author Declarations

Ethical oversight of the project was provided by Public Health Wales (PHW) Research & Development (R&D) Division. The PHW R&D Division advised that NHS research ethics approval was not required as this work was classed as service evaluation as it met the HRA criteria for post market surveillance studies of CE marked devices. Data were held and processed under Public Health Wales information governance arrangements, in compliance with the Data Protection Act, Caldicott Principles and Public Health Wales guidance on the release of small numbers. No data identifying protected characteristics of an individual were released outside Public Health Wales. PHW R&D Office is both an entry point and sub-department of NHS research ethics board.

